# Implementation and Evaluation of Obstetric Early Warning Systems in tertiary care hospitals in Nigeria

**DOI:** 10.1101/2020.09.10.20166140

**Authors:** Aminu Umar, Saidu Ibrahim, Idris Liman, Calvin Chama, Munirdeen Ijaiya, Matthews Mathai, Charles Ameh

## Abstract

**Background:** Obstetric Early Warning Systems (EWS) use combined clinical observations to predict increased risk of deterioration and alert health workers to institute actions likely to improve outcomes. The objective of this study was to explore the experience of health workers/managers who implemented a low resource setting-specific statistically derived and validated EWS and to assess its effectiveness in improving health outcomes.

**Methods:** This mixed-method study included 2400 women admitted to inpatient wards between 1 August 2018 and 31 March 2019 at three tertiary Nigerian hospitals (1 intervention and 2 control) with pregnancy and childbirth-related complications. The quality of patient monitoring and prevalence of outcomes were assessed through retrospective review of case notes before and 4 months after EWS was introduced. Outcomes were maternal death, direct obstetric complications, length of hospital stay, speed of clinical review, caesarean section(CS) and instrumental birth rates. Qualitative interviews and focus group discussions were undertaken to explore the views of healthcare workers on EWS’ acceptability and usability.

**Results:** EWS was correctly used in 51% (n=307) of cases. Of these, 58.6% (180) were predicted to have increased risk of deterioration, and 38.9% (n=70) were reviewed within 1 hour. There was a significant improvement in the frequency of vital signs recording in the intervention site: observed/expected frequency improved to 0.91 from 0.57, p<0.005, but not in the control sites. CS rate dropped from 39.9% to 31.5% (chi-square p=0.002). No statistically significant effect was observed in the other outcomes.

Health workers reported positive experience using EWS, with the feeling that it helped cope with work demands while making it easier to detect and manage deteriorating patients. Nurses and doctors reported that the EWS was easy to use, evaluate at a glance, and that scores consistently correlated with the clinical picture of patients. Identified challenges to use included rotation of clinical staff, low staffing numbers and monitoring equipment.

**Conclusion:** The implementation of EWS improved the quality of patient monitoring, but a larger study will be required to explore the effect on critical care admission and health outcomes. With modifications to suit the setting, coupled with regular training, the EWS is a feasible and acceptable tool to cope with the unique demands faced in low-resource settings.

Summary box
What is already known?

- An obstetric EWS algorithm with seven parameters (RR, temperature, systolic BP, pulse rate, consciousness level, urinary output and mode of birth), was developed and internally validated using data from low resource settings.

What are the new findings?

- There was a significant improvement in the frequency of vital signs recording following implementation of the algorithm in the intervention site: observed/expected frequency improved to 0.91 from 0.57, p<0.005, but not in the control sites.
- CS rate dropped from 39.9% to 31.5% (chi-square p=0.002) in the intervention arm, though difficult to attribute to reduction in medically unnecessary CS.
- The obstetric EWS algorithm was easy to use, easy to evaluate at a glance, capable of guiding referral and accurate, with scores always correlating with the clinical picture of patients.

What do the new findings imply?

- EWS is feasible to implement and potentially acceptable tool to cope with the unique demands faced by obstetric practice in low-resource settings

## Introduction

Although the global maternal death burden has fallen by almost 50 per cent in the last two decades, still an estimated 810 deaths occur daily around the world due to complications of pregnancy and childbirth (WHO, U., 2019). Most of these deaths (94%) occurred in low-resource settings, and most could have been prevented (WHO, U., 2019). It is also estimated that there are 27 million episodes of direct obstetric complications annually which contribute to long-term pregnancy and childbirth complications (Machiyama et al., 2017).

Nigeria accounted for 20% of the reported global maternal deaths in 2015 (WHO Factsheet, 2016) (WHO, 2016). Marked inequalities exist, with northern regions having significantly higher maternal deaths compared to the southern regions (Meh C, et al 2019).

Increased facility-based births has been reported in the last 15 years, in all WHO regions, with the proportion of birth, attended by skilled health personnel rising from 56% in 1990 to almost 80% by 2017 (United Nations, 2018). With the resulting increase in the utilisation of health services, a higher proportion of preventable maternal morbidity and mortality has moved from communities to health facilities. Consequently, opportunities to ensure good quality facility care, including timely diagnosis and management of obstetric complications are critical, if the new ambitious maternal health targets of the Sustainable Development Goals (SDG) are to be achieved (United Nations, 2019).

Early Warning Systems (EWS) have been developed to facilitate the timely presence of appropriately skilled staff to attend clinically deteriorating patients (Morgan, 2007). They provide the opportunity to aggregate the impact of sometimes subtle deteriorations in physiological observations into an overall score that, when abnormal, is used to prompt a clinical response (Alam et al, 2014). However, the early warning systems designed for the general population does not account for the unique physiology of pregnant women, and it does not effectively identify at-risk obstetric patients (Lappen JR et al, 2010).

Obstetric EWS is recommended for monitoring the condition of hospitalised pregnant and postnatal women, based on predetermined abnormal values (warning signs) to generate a rapid medical response and facilitate early detection and management of clinical deterioration (ACOG, 2013; CEMACH, 2007; Edwards et al., 2015; Mhyre et al., 2014; Shields, 2016). A recent systematic review of EWS used in obstetrics reported that they are effective in predicting adverse obstetric outcomes and reducing obstetric morbidity (Umar A. et al, 2019).

Several EWS have been developed for obstetric patients, but the majority are the result of a clinical consensus rather than formal statistical analyses or were created using data from patients admitted to intensive care units, limiting their generalizability to non-intensive care settings (Carle et al, 2013; Hedriana, et al, 2016; Isaacs et al., 2014; Mhyre et al., 2014; Paternina-Caicedo et al., 2017; Shields, 2016; Singh S, et al, 2012). Using secondary data on obstetric inpatients admitted to 42 Nigerian tertiary hospitals, Umar A. et al (2020) developed and internally validated a simple obstetric diagnostic prediction model and EWS for use in resource-limited settings using recommended methodologies (Umar A., et al 2020; Collins, et al, 2015). The resulting EWS model performed excellently in predicting Severe Maternal Outcome (SMO: maternal death or near-miss) in the derivation data set with AUROC consistently above 90% and demonstrated a potential for usefulness in other similar settings (Umar A., et al 2020). The objectives of this study were to explore the experience of health workers about the implementation of this EWS, and to assess its effectiveness in improving health outcomes.

## Materials and methods

### Hypothesis

This study tested the hypothesis that the statistically developed EWS reported by Umar A. et al (2020) will perform equally well in a different setting than its derivation population. We also hypothesised that EWS chart will potentially provide an easier, more convenient and efficient alternative clinical monitoring method than the routine practice.

### Study design

A mixed-method research design consisting of a controlled before-after quasi-experimental trial, qualitative interviews and focus group discussions were employed to achieve the study objectives. The study was conducted over an 8-month period (between 1 August 2018 and 31 March 2019).

### Setting

The study was implemented in 3 tertiary care hospitals across the northern regions of Nigeria. The EWS was implemented in the 600-bed multispecialty University of Ilorin Teaching Hospital, a public tertiary health care centre located in the north-central region. The obstetric and gynaecology unit of the hospital had 208 bed capacity for obstetric and gynaecology cases managed by 4 teams of consultans, registrars and medical interns. The control sites, National Hospital Abuja (control 1), and Abubakar Tafawa Balewa university teaching hospital Bauchi (control 2) are teaching hospitals in the north-central and northeast regions, respectively.

### Participants and recruitment

Pregnant and postpartum women admitted to all inpatient wards due to complications developing antepartum or during the puerperium (42 days’ post-partum) were included in the study. Women were excluded if they were in active labour, were discharged within 24 hours of normal vaginal birth, or had met any of the three maternal near-miss criteria before hospital admission (clinical, management-based and organ dysfunction-based criteria (Say, Souza, & Pattinson, 2009)). Women who were admitted directly to intensive care unit without going through any of the inpatient wards were also excluded. Recruitment was conducted by trained nurses and midwives undertaking clinical monitoring in the respective wards.

### Intervention

The intervention was the use of a statistically developed obstetric EWS, details of which were published elsewhere (Umar A., et al, 2020). The resulting EWS chart (***Annex 1****: EWS chart*) was introduced to replace the vital signs charts of all recruited participants in the intervention site. Briefly, this is a simple score-based recording chart for vital signs. It includes seven clinical parameters (temperature, pulse rate, respiratory rate, systolic blood pressure, diastolic blood pressure, consciousness level (based on the AVPU (alert, voice, pain and unresponsive) scale) and mode of birth for post-partum women). Each parameter is scored as 0 for normal, 1 for mild and 2 for severe derangements. An escalation protocol at the top of the chart guides frequency of patient monitoring and when to trigger clinicians’ review (***Annex 1****: EWS chart*); scores of 0 or 1 are reassuring; hence require 12-hourly monitoring or as routine for post-operative patients. A score of 2 indicates the need to repeat observations after 30 minutes; if the score remains the same or rises, doctors should be informed for review. Those with scores of 3 or more are likely to deteriorate clinically and require immediate review. The two control sites were asked to continue with the existing practices of clinical monitoring.

### Outcome measures

Primary outcomes assessed were maternal death and direct obstetric complications (pre-eclampsia/ eclampsia, antepartum haemorrhage, postpartum haemorrhage, sepsis, prolonged/obstructed labour, abortions complications, and thromboembolism). Secondary outcome measures considered included the frequency of vital signs monitoring and recording. This was assessed using the patient monitoring index (PMI), defined as the ratio of the observed to the expected frequency of vital signs monitoring over 24 hours. Others were duration of hospital stay, speed of post-EWS trigger specialist review, caesarean section rate and rate of instrumental delivery.

### Sample size calculation

Preliminary data from the intervention site showed that the average monthly obstetric admission was 190 patients. Of these, about 28% experienced a maternal death or direct obstetric complications (haemorrhage, sepsis, abortion complications, hypertensive disorders, prolonged/obstructed labour and thromboembolism).

Sample size estimate was made using a baseline outcome prevalence of 25-30% as illustrated in Table 1. We considered a three-month period for each of the pre- and post-intervention follow-ups as further increase does not significantly improve the power (Table 1). Factoring in a possible exclusion rate of around 5%, the sample size considered was 1200 in the intervention site (600 each pre- and post-intervention). The same numbers of participants were to be recruited in the control sites (Table 1).

**Table 1:** Sample size calculation

| Number in pre-intervention | Number in the follow-up period | Baseline of 25% |  | Baseline of 30% |  |
| --- | --- | --- | --- | --- | --- |
|  |  | Reduction detectable |  | Reduction detectable |  |
|  |  | 80% power | 90% power | 80% power | 90% power |
| 380 (2 months) | 570 (3 months) | 8.0% | 9.1% | 8.6% | 9.8% |
|  | 760 (4 months) | 7.6% | 8.7% | 8.1% | 9.3% |
|  | 950 (5 months) | 7.3% | 8.4% | 7.8% | 9.0% |
|  | 1140 (6 months) | 7.1% | 8.2% | 7.6% | 8.8% |
| 570 (3 months) | 570 (3 months) | 7.2% | 8.2% | 7.7% | 8.8% |
| Sample size by study arms |  |  |  |  |  |
| Study arm | Before | After |  | Total |  |
| Intervention | 600 | 600 |  | 1200 |  |
| Control 1 | 300 | 300 |  | 600 |  |
| Control 2 | 300 | 300 |  | 600 |  |
| Total | 1200 | 1200 |  | 2400 |  |

Intervention = UITH Ilorin, Control 1 = National hospital Abuja, Control 2 = ATBUTH Bauchi

## Trial procedure

Ethical approvals were received from the Research and Ethics Committee of the Liverpool School of Tropical Medicine (LSTM-Research Protocol 18-074) and the three study sites (NHA/OG/GC/0171, UITH/CAT/189/19/167, ATBUTH/ ADM /42 / VOL1). Since routinely collected patient information was used for the research, no individual-level consent was deemed necessary as was performed in similar studies (Merriel et al., 2017; Sheikh et al., 2017; Singh, A., et al., 2016).

Training workshops on EWS were conducted in the intervention hospital in November 2018. Following this, the hospital management updated the hospital’s guideline for patient monitoring. Specifically, the EWS was to replace routine vital signs charts for all obstetric inpatients (emergency, antenatal, postnatal medical, postnatal surgical and gynaecology wards). A local implementation team consisting of doctors and nurses was constituted to support implementation. They provided on-the-job training on-demand regarding the use of the EWS, including the use of an escalation protocol demonstrated through the use of practical case scenarios. The team also responded to any queries about the implementation of the EWS. Management of specific complications was as per hospital protocol or usual practice as appropriate.

Compliance was audited by the local implementation team daily until when all obstetric inpatients had the EWS in their case notes, at which point recruitment began. Thereafter, weekly audits were conducted to ensure that the charts were correctly used on all participants. Additionally, formal monthly audits were performed by the principal researcher on a randomly selected date to monitor the use of the chart and any ongoing change in practice. A quality indicator was adapted from that described by Merriel et al (2017) to provide the implementation team with an easy, but an objective way to assess the quality of the implementation. (**Annex 2:** Quality audit checklist) (Merriel et al., 2017) This indicator was used to audit the EWS charts of all obstetric inpatients on the day of the audit. The quality indicator measures three essential ratios: 1) the usage rate of charts (number of patients with correctly completed EWS charts/number of charts reviewed), 2) whether healthcare staff took appropriate action on abnormal observations (number of cases in which action was taken/total number of charts requiring action), and 3) the timeliness of the action if one was required (total number where the action was taken within the required time frame/total number where the action was taken).

## Quantitative data collection and management

Baseline retrospective case note reviews of 1200 obstetric admissions to the three health facilities between 1 August and 31 October 2018 was conducted by 3 research assistants. Android data collection devices (ODK Collect) were used for all quantitative data collection.

Following this, the EWS chart was implemented in the intervention hospital in November 2018. A transition period of two weeks was allowed to audit implementation before the recruitment commenced for prospective post-implementation data. Following this, women were recruited into the study on admission to all five obstetric inpatient wards by the ward nurses, and EWS were incorporated into their medical files. They were prospectively followed until the end of the stay in hospital (discharge or demise) during which a dedicated research assistant (a medical intern) retrieved all completed EWS charts for analysis daily. Women in active labour were excluded and monitored with partograph as defined by the study protocol. Data were collected in the two study arms for four months after implementation (1 December 2018 to 31 March 31 2019) until the desired sample size (n=600) was achieved.

These data were exported to a Microsoft Excel spreadsheet and cleaned for analysis. Data analysis was conducted using IBM SPSS version 23. Normality of distribution of variables was assessed by using distribution plots and Shapiro-Wilk testing. Cumulative and facility-specific characteristics were summarized by mean (SD) for continuous variables and percentages for categorical variables. Descriptive statistics were calculated.

Prevalence of outcomes during the post-implementation period at the control sites was reported to check for trend effects or effects of other quality improvement programmes during the study period. Outcome measures were compared within and between study arms using independent sample t-testing and chi-square for continuous and categorical variables respectively. *P<*0.05 was considered as statistically significant.

## Qualitative data collection and management

Semi-structured key informant interviews (KII) and focus group discussions (FGD) were conducted in the intervention hospital at the end of the follow-up period in April 2019 to explore the experience of health workers in the use of the EWS.

The KIIs participants (n=12) were purposively selected senior midwives/nurses in administrative positions and doctors in the Obstetrics department. The FGDs (n=6) were conducted with junior nurses/midwives who undertake monitoring of obstetric patients using the EWS. Through the FGD, we aimed to explore and understand their experience of implementing the EWS.

Open-ended interview questions were predesigned based on the study objectives and emerging themes during the post-implementation data collection. Interviews and FGDs conducted in English language, audio recorded and transcribed verbatim. Data was collected until saturation, and analysed using thematic framework approach (Richie, J., Spencer, L., 2003).

## Results

### Recruitment

Overall, 4258 women were admitted to the three hospitals for childbirth or pregnancy complications between 1 August 2018 and 31 March 2019. A total of 3997 live births and 273 stillbirths were recorded, placing the overall SBR at 63.9/1000 births. Nearly one in five births was preterm (before 37 completed weeks of gestation).

During the baseline pre-implementation period (1 August to 31 October 2018), 1200 women were recruited into the study retrospectively, comprising 600 in the intervention site, and 300 each in the two control sites. Following surveillance, a target of 100% implementation of EWS among all obstetric admissions to five inpatient wards was achieved by the end of November 2018.

Recruitment started from 1 December 2018 in the intervention arm, during which the highest recruitment rate of 95.2% was reported. However, the recruitment rate fell significantly in January 2019 when only 70.8% of research-eligible patients were recruited into the study, but rose steadily thereafter, reaching a peak of 78.1% by the end of March 2019. Overall, the required sample size (n=600) was achieved after four months (1 December 2018 to 31 March 2019), with an average recruitment rate of 78.8% (**Figure 1**).

**Figure 1.**
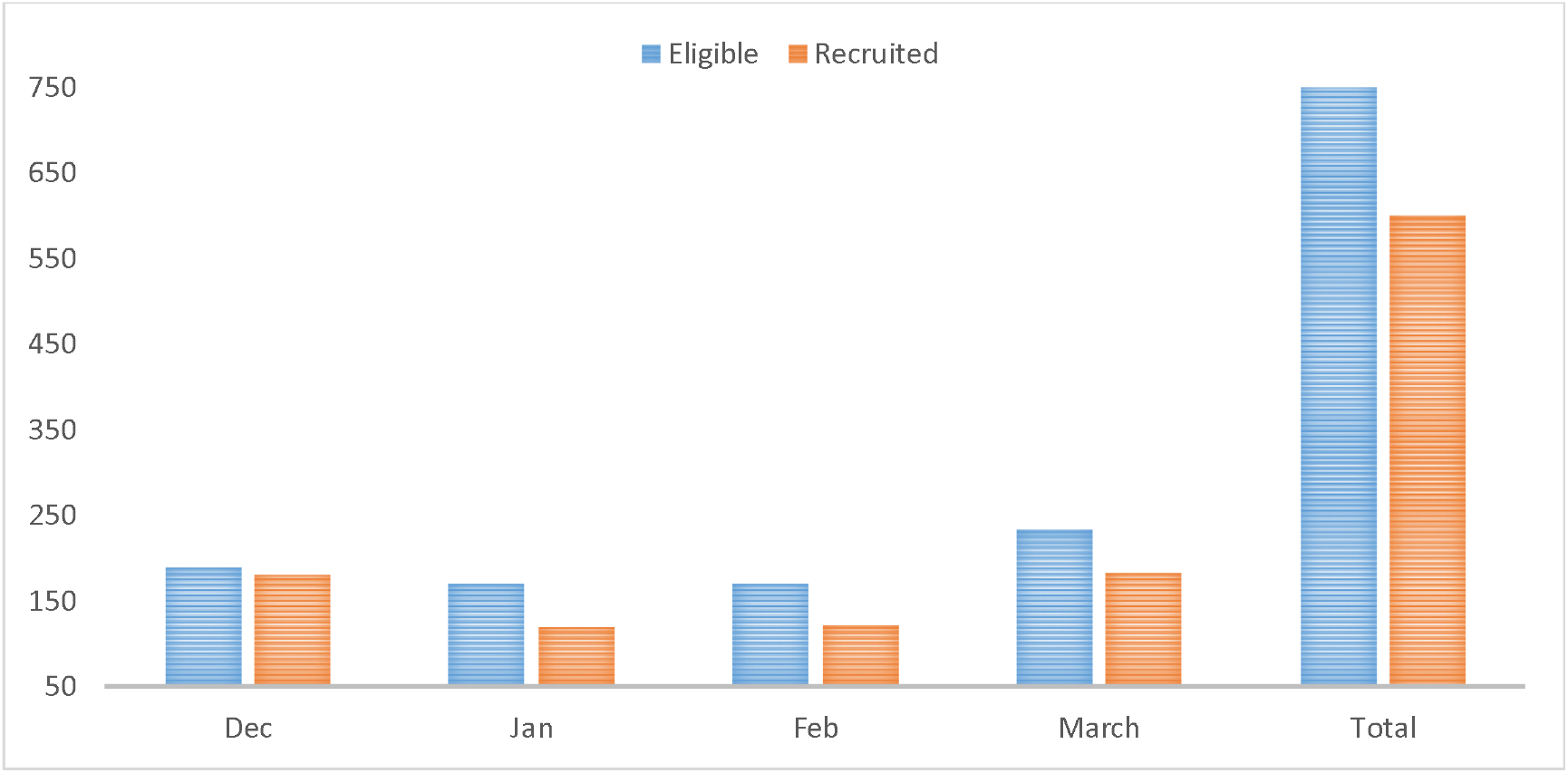
Recruitment of participants in the intervention site (1 December 2018 to 31 March 2019)

## Baseline characteristics

The characteristics of the women by study arms are illustrated in **Table 2**. There was no difference in age between the intervention and control groups at baseline (p=0.348) and post implementation (p=0.169). More women were registered for antenatal care in the control hospitals at baseline (p=0.024); however, this difference ceased to be significant in the post-implementation cohort (p=0.155) (**Table 2**).

**Table 2.** Characteristics of study participants

| Characteristic | Baseline (n=1200) |  |  | Post-implementation (n=600) |  |  |
| --- | --- | --- | --- | --- | --- | --- |
|  | Intervention (n=600) | Control (n=600) | P-value | Intervention (N=600) | Control (n=600) | P-value |
| Age (years) | 30.0 (5.3) | 28 (6.4) | 0.348 | 30 (5.2) | 28 (6.3) | 0.169 |
| Weight (kg) | 72.0 (14.4) | 63 (14.2) | **0.038** | 70 (11.7) | 62.7 (13) | **0.023** |
| Height (m) | 1.6 (0.05) | 1.6 (0.1) | 0.673 | 1.6 (0.06) | 1.6 (.09) | 0.334 |
| LOS (days) | 3.6 (3.2) | 2.4 (1.8) | 0.368 | 3.7 (3.5) | 2.3 (1.6) | 0.714 |
| Booked (%) | 60.8 | 72.2 | **0.024** | 65.7 | 68.4 | 0.155 |
| Booking GA (days) | 24.8 (8.4) | 25.5 (8.1) | 0.906 | 24.6 (8.4) | 25.1(8.1) | 0.531 |
| ANC visits | 2.6 (1.3) | 4.3 (2.3) | **0.036** | 2.7 (1.7) | 4.4 (2.3) | **0.042** |
| Parity | 2.2 (1.4) | 2.9 (2.3) | 0.305 | 2.2 (1.3) | 3.0 (2.3) | 0.181 |

| Obstetric complications (%) | Baseline (n=1200) |  |  | Post-implementation (n=600) |  |  |
| --- | --- | --- | --- | --- | --- | --- |
|  | Intervention (n=600) | Control (n=600) | Chi-sq. P-value | Intervention (N=600) | Control (n=600) | Chi-sq. P-value |
| Haemorrhage | 10.7 | 4.2 | **0.027** | 11.1 | 4.9 | **0.019** |
| Sepsis | 12.2 | 8.8 | **0.044** | 12.7 | 8.7 | **0.040** |
| Hypertensive disorders | 10.5 | 7.5 | 0.125 | 9.8 | 7.6 | 0.331 |
| Prolonged labour | 10.3 | 7.7 | 0.472 | 10.4 | 8.0 | 0.533 |
| Obstructed labour | 9.8 | 4.3 | **0.018** | 9.8 | 5.8 | **0.049** |
| Thromboembolism | 0.3 | 0.0 | NA | 0.2 | 0.0 | NA |
| Abortions | 5.8 | 2.5 | **0.023** | 6.4 | 2.4 | **0.040** |

LOS: Length of hospital stay; Booked- patients registered for ANC; Booking GA- Gestational age at ANC registration

Fifty women died due to causes related to pregnancy or childbirth across the three health facilities, putting the cumulative estimated MMR at 1052 per 100,000 live births. Facility-level estimates showed a similar prevalence of maternal death in the intervention site and control hospital-2 (institutional MMR of 1393 and 1320 per 100,000 live births respectively), both having over three times as many deaths as control hospital-1 (institutional MMR of 440 per 100 000 live births).

Overall, maternal morbidity rate was higher in the intervention hospital. During the pre-implementation period, twice as many women suffered obstetric haemorrhage in the intervention hospital compared to the controls. Similarly, the prevalence of obstructed labour and abortions in the intervention arm was twice that of the control hospitals. The commonest obstetric complication was sepsis, which complicated 12.2% and 8.8% of the obstetric admissions in intervention and control hospitals respectively. Although prevalence of both hypertensive disorders and prolonged labour were higher in the intervention site, the difference was not statistically significant nor was the difference in ICU admission rates (**Table 2**).

Similar distribution of maternal morbidity was seen across study arms in the post-implementation period (**Table 2**). Women were nearly three times more likely to suffer obstetric haemorrhage or abortions (16.5%), and twice as likely to have obstructed labour (9.8%) in the intervention hospital compared to the controls (6.7% and 4.3%, respectively). Sepsis also remained the commonest complication, affecting 12.7% and 8.7% of obstetric admissions in the intervention and control hospitals respectively (**Table 2**).

## Completion and trigger rate of EWS

Overall, recording of EWS parameters was incomplete, with regular monitoring (at least twice in 24 hours) of temperature, pulse, respiratory rate and blood pressure performed in 54% of the study participants (**Figure 2**). Most patients (over 89.2%) had all vital signs monitored and recorded at least once in 24 hours.

**Figure 2.**
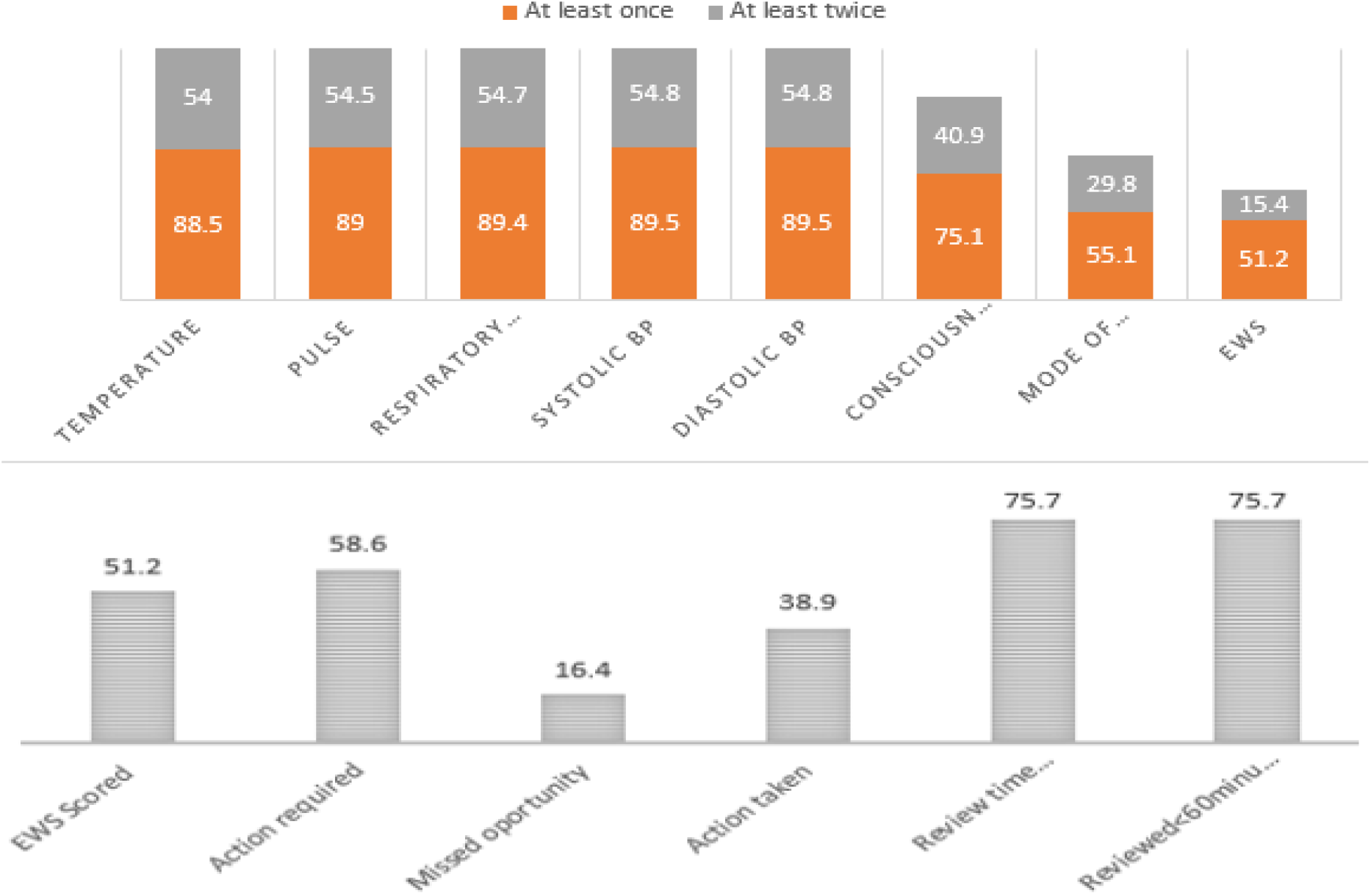
Completion rate of the EWS parameters and trigger system

Although monitored and recorded, EWS parameters were converted and summed into an EWS score in significantly fewer patients; only 15.4% (n=92) of the study participants had EWS scores documented as prescribed by the study protocol (at least twice in 24 hours). About half of the study participants (51.2%, n=307) had EWS scores recorded at least once in 24 hours (**Figure 2**).

About 58.6% (n=180) of the 307 women who had EWS score documented at least once in 24 hours required medical review by a doctor (**Figure 2**). Of these, 38.9% (n=70) were reviewed by a doctor. In terms of timeliness of the review, about three-quarters of the reviewed patients (75.7%, n=53) had the time of doctor’s review correctly documented on the EWS chart; all of these patients were reviewed within 60 minutes, as recommended by the EWS escalation protocol.

## Analysis of outcomes

There was no significant difference in maternal mortality rate between pre-implementation and post-implementation phases in both two arms of the study (**Table 3**). No maternal near misses based on WHO near-miss criteria were recorded in any of the three hospitals. Hence, maternal morbidity was defined as diagnosed by clinicians in the patients’ medical records. Although the prevalence of morbidity varied significantly across the study arms (**Table 2**), there was no change in prevalence within the trial arms following EWS implementation (**Table 3**). Similarly, there was no change in the length of hospital stay and ICU admission rates.

Caesarean section rate dropped significantly from 39.9% during the baseline period to 31.5% following implementation of the EWS in the intervention hospital. A significant rise was observed in the average CS rate of the two control hospitals (**Table 3**) in the post-implementation period. However, a sensitivity analysis showed a disproportionately higher caesarean birth rate in control site 1 compared to 2 (61.4% and 22.1% respectively). Hence, a facility-level analysis was performed, which showed no significant change in the caesarean section rate in the two control hospitals during the post-implementation period from the baseline rates.

Overall instrumental delivery rate was very low in all three hospitals. Only four forceps deliveries were reported throughout, all conducted in control site 2. No instrumental births were performed in control site 1, hence the estimate used in the outcome analysis was derived using data from control site 2 (**Table 3**). There was no significant change (by Fisher’s exact test) in the rate of vacuum deliveries following EWS implementation in the intervention hospital. Similarly, no significant change in the rate of instrumental deliveries (vacuum and forceps births) was observed in the control arm (**Table 3**).

**Table 3:**
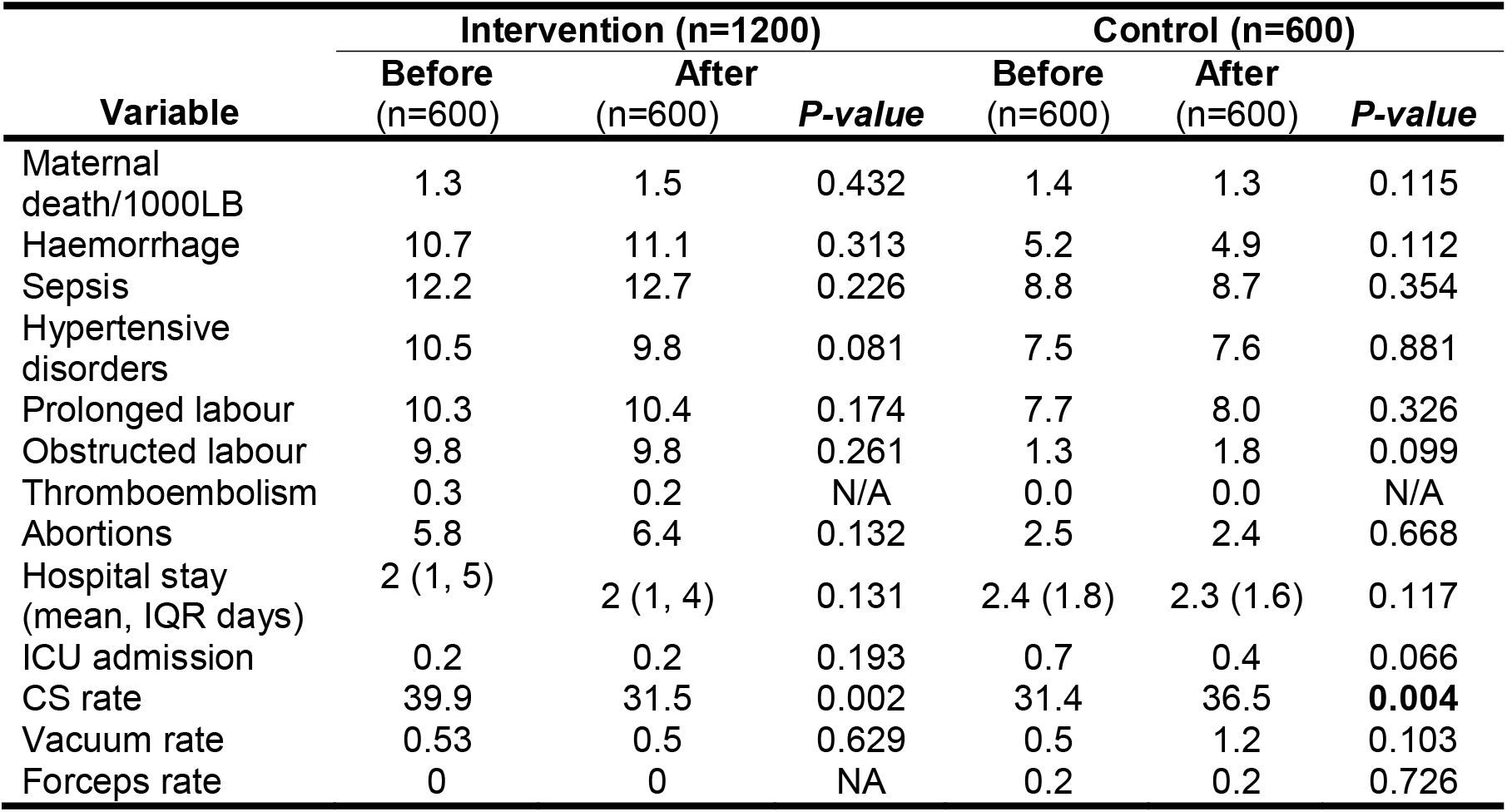
Prevalence of outcome measures before and after EWS implementation

| Variable | Intervention (n=1200) |  |  | Control (n=600) |  |  |
| --- | --- | --- | --- | --- | --- | --- |
|  | Before<br>(n=600) | After<br>(n=600) | *P-value* | Before<br>(n=600) | After<br>(n=600) | *P-value* |
| Maternal death/1000LB | 1.3 | 1.5 | 0.432 | 1.4 | 1.3 | 0.115 |
| Haemorrhage | 10.7 | 11.1 | 0.313 | 5.2 | 4.9 | 0.112 |
| Sepsis | 12.2 | 12.7 | 0.226 | 8.8 | 8.7 | 0.354 |
| Hypertensive disorders | 10.5 | 9.8 | 0.081 | 7.5 | 7.6 | 0.881 |
| Prolonged labour | 10.3 | 10.4 | 0.174 | 7.7 | 8.0 | 0.326 |
| Obstructed labour | 9.8 | 9.8 | 0.261 | 1.3 | 1.8 | 0.099 |
| Thromboembolism | 0.3 | 0.2 | N/A | 0.0 | 0.0 | N/A |
| Abortions | 5.8 | 6.4 | 0.132 | 2.5 | 2.4 | 0.668 |
| Hospital stay (mean, IQR days) | 2 (1, 5) | 2 (1, 4) | 0.131 | 2.4 (1.8) | 2.3 (1.6) | 0.117 |
| ICU admission | 0.2 | 0.2 | 0.193 | 0.7 | 0.4 | 0.066 |
| CS rate | 39.9 | 31.5 | 0.002 | 31.4 | 36.5 | **0.004** |
| Vacuum rate | 0.53 | 0.5 | 0.629 | 0.5 | 1.2 | 0.103 |
| Forceps rate | 0 | 0 | NA | 0.2 | 0.2 | 0.726 |

Frequency of monitoring of patients was assessed using PMI for the four routinely monitored vital signs (respiratory rate, temperature, pulse rate and blood pressure). Across all three hospitals, the guidelines for monitoring obstetric patients using the vital signs chart is to monitor them every 6 hours (at least 4 times in 24 hours). While this applied for the intervention hospital during the baseline period, the expected frequency of monitoring during the post-implementation period was as specified by the EWS escalation protocol; i.e. twice daily for EWS scores of 0 or 1, 30-minutes apart for a score of 2 and immediate referral for scores of 3 or more.

**Table 4.** Frequency of vital signs monitoring before and after EWS implementation Intervention Hospital: Patient Monitoring Indices (PMI)

| **Intervention Hospital: Patient Monitoring Indices (PMI)** |  |  |  |
| --- | --- | --- | --- |
|  | **Baseline** | **After** | **t-test (p)** |
| Temp (SD) | 0.5 (0.4) | 0.9 (0.4) | <0.005 |
| Pulse (SD) | 0.6 (0.3) | 0.9 (0.4) | <0.005 |
| RR (SD) | 0.5 (0.4) | 0.9 (0.4) | <0.005 |
| BP (SD) | 0.6 (0.3) | 0.9 (0.4) | <0.005 |
| **Control Hospitals: Absolute monitoring frequencies** |  |  |  |
|  | **Baseline** | **After** | **t-test (p)** |
| Temp (SD) | 1.7 (0.9) | 1.6 (0.9) | 0.234 |
| Pulse (SD) | 1.8 (0.9) | 1.7 (0.9) | 0.123 |
| RR (SD) | 1.8 (1.3) | 1.8 (1.2) | 0.221 |
| BP (SD) | 1.8 (0.9) | 1.7 (0.9) | 0.115 |

Significant improvement in the frequency of monitoring was observed in the intervention hospital (**Table 6.4**). This was especially so for temperature and respiratory rate monitoring, with baseline mean (SD) PMI of 0.5 (0.4) and 0.5 (0.4), and post-implementation mean (SD) PMI of 0.9 (0.4) and 0.9 (0.4) respectively. No significant change in the frequency of vital signs monitoring was observed in both control hospitals (**Table 4**).

## Experience and challenges of using EWS

Most of the nurses/midwives found the EWS chart useful in alerting them when to escalate care to doctors. They reported that abnormal observations are usually an indicator that the patient needs more frequent monitoring. In addition to contributing to the early detection of deterioration, they felt the chart assisted them directly in managing sick patients.

Compared to the routinely used vital signs chart, most of the nurses felt EWS was easier to use because of less frequent monitoring of clinically stable patients. By scoring vital signs and having a cumulative EWS score, the chart “*compresses clinically relevant parameters into a simple score, making it easy to evaluate patients at a glance* (*KII nurse*)”.

The doctors opined that EWS was a good monitoring tool if properly followed. They found the charts easy to correlate with a patient’s clinical picture, with abnormal scores usually consistent with clinical deterioration. They also felt the chart could potentially help nurses to cope with the demands of their work, given the gross shortage of human resource for health, while making it easier to detect unwell patients.

Overall, most interviewees agreed a colour-coded EWS would be easier to use and more efficient in picking out and communicating the need for clinical review. Additionally, it would be less labour-intensive and more visually appealing, hence more likely to be accepted by clinical staff.

Major limiting factors to effective monitoring of vital signs using EWS were the shortage of functioning equipment and frequent staff rotation by the hospital management. There was a gross shortage of patient monitoring equipment across all five wards. Although the hospital management had approved the use of the EWS instead of the routinely used vital signs charts, some nurses reported having to use the old monitoring chart concurrently with the EWS charts, potentially increasing the work load of staff and stretching the use of scarce patient monitoring equipment.

Rotations of nurses/midwives (and medical interns), that happen every 6-months, brings in new clinical staff who are untrained in the use of EWS. This happened shortly after the EWS implementation, taking most of the trained nurses to other clinical departments. This significantly affected the recruitment rate and overall success of the study. A few midwives reported that the escalation protocol was ambiguous, hence a common cause of error in patient monitoring, especially among newly deployed staff nurses. The training provided was said to be grossly inadequate.

Shortages in human resources, especially of staff nurses and midwives were also reported as a major challenge. Afternoon shifts in some wards were covered by only one staff nurse who looked after at least 10 patients. Delayed supplies of the EWS had made some wards resort to using the old vital signs charts when EWS ran out. A few nurses also reported having had to use the old chart concurrently with the EWS because the latter was always retrieved at the end of hospital stay for analysis. Selected quotes on box 1.0 illustrate the above findings.

#### >Box 1.0: Staff comments on the EWS

*Clinical information of patients is compressed into a single score, making it easy to evaluate at a glance … (FGD nurse)*

*it was accurate in that all patients with high scores are always the sick ones. In fact, it even assists us in monitoring how our post-operative patients are recovering after caesarean section… (KII Nurse)*

*You know interns rotate, so are nurses, most of the errors in scoring are caused by lack of continuous training (KII Doctor)*

*we don’t have enough staff, like in the postnatal medical ward, you find only one nurse covering most afternoon shifts… (KII nurse)*

*like here we have only one BP machine on the ward, and to check patient’s oxygen saturation you have to transfer her to gynaecology emergency ward… (FGD nurse)*

*A coloured chart would be easier to use than this… am sure you chose black and white because of low production cost, but the management should adopt the coloured chart… (KII Doctor)*

## Discussion

Although a lot of work has taken place to assess the benefits of obstetric EWS across the globe, to the best our knowledge, this is the first report of the implementation of EWS in a low resource tertiary hospital, that was statistically derived and internally validated using data from low resource settings.

The average institutional maternal mortality ratio across the 3 study sites was 1052 per 100,000 live births. This is significantly higher than the population-based national average of 917 per 100,000 live births (WHO, U., 2019), but comparable to an institutional-based estimate of 1088 per 100,000 (Oladapo et al., 2015). We observed no change in MMR following EWS implementation. Maternal mortality reviews indicate that a significant proportion of women who die due to pregnancy and childbirth demonstrate abnormal vital signs long before death, suggesting that a multi-parameter EWS system should identify them with high specificity (Mhyre et al., 2014b) (Carle et al., 2013; Paternina-Caicedo et al., 2017). Effectively, this should facilitate timely diagnosis and treatment, limit the severity of morbidity, and possibly reduce mortality. Although our findings are consistent with the systematic review report of limited evidence on the effectiveness of obstetric EWS in reducing maternal death (Umar A., et al., 2019), maternal mortality is an extremely rare outcome; hence, the considerably large sample size is required to have a substantial number for effective analysis. Given that the implementation of EWS in this study involved a single facility with only 18 maternal deaths over the seven months of the research, large multicentre randomized controlled trials will be required to evaluate the effectiveness of obstetric EWS in reducing death.

There was no change in the proportion of obstetric complications, ICU admission and length of hospital stay following EWS implementation. Obstetric EWS have been previously shown to prevent progressive obstetric morbidity (Shields et al., 2016; Hedriana et al., 2016; Umar A., et al., 2019). In contrast to our findings also, Shields and colleagues, in a large multicentre quasi-experimental trial, reported a significant reduction in severe and composite maternal morbidity (p<0.01) as defined by the Centre for Disease Control (CDC), but not mortality, in six intervention hospitals following EWS implementation, compared to 19 controls (Shields et al., 2016). However, they also observed no change in the ICU admission rate in either the intervention or control hospitals (Shields et al., 2016). We are aware that clinical outcomes would only have been improved if completion of the EWS had led to an escalation of intervention, physician assessment as specified by the system, and the institution of appropriate interventions. It could be argued that the period post-implementation (four months) would not have been sufficient to allow such changes to manifest, especially considering the designs of similar studies that measured outcomes after longer periods (12–18 months) (Merriel et al., 2017; Moore et al., 2019; Shields et al., 2016). It could also be argued that the lack of standardization of morbidity outcomes used in our analysis limits its comparability to findings of similar studies. The latter limitation is, however, not specific to our research, as previously reported in EWS validation studies (Singh, A., et al., 2016; Singh, S., et al., 2012; Umar A., et al., 2019).

The patient monitoring index showed a significant improvement following EWS implementation in the intervention hospital. This was especially so for temperature and respiratory rate, but also significant for the remaining EWS parameters. With a similar baseline frequency of vital signs monitoring as the intervention hospital, no change in frequency was observed in the control hospitals during the post-implementation period. This is consistent with findings of one before-after study, which reported an increase in the frequency of documentation of vital signs following the implementation of the Irish Maternity EWS on obstetric patients with sepsis (Maguire, et al., 2015). Our findings also support the finding that introduction of modified obstetric EWS improves the post-operative monitoring of women after caesarean section (Merriel et al., 2017; Moore et al., 2019; Sheikh et al., 2017).

Caesarean births constituted 31.8% of all births in the 3 hospitals. This is a significant increase from a 2009 report of hospital-level CS rate of 14% in Nigerian tertiary hospitals (Shah et al., 2009). in the last three decades, the caesarean section rate has continued to increase in an unprecedented manner in both developed and developing countries, above the WHO recommended 10% at population level (Boerma et al., 2018; WHO, 2015). This rise is driven by major increases in non-medically indicated CS (Betran et al., 2015; Vela et al., 2014). Following the implementation of EWS, the CS rate dropped significantly in the intervention, but not in any of the control facilities. However, it was difficult to attribute this to a reduction in medically unnecessary CS as an individual-level analysis of indications for caesarean births during the baseline and post-implementation periods was not performed.

The instrumental delivery rate was very low and remained unchanged following EWS implementation. This is not peculiar to our study settings, as there has been a decline in the reported rates worldwide (Okeke & Ekwuazi, 2013). The rates vary greatly between settings and the ideal is unknown (Ameh, C., et al., 2009). With the caesarean section rate known to vary inversely as the rate of instrumental delivery (Dildy et al., 2016), a high CS rate could have played a key role in the observed low uptake in our study cohort. Nevertheless, discussing the implications of low uptake of forceps and vacuum deliveries is outside the scope of this paper.

The opinion of clinical staff regarding EWS was generally positive, with the feeling that it helped them to cope with the demands of their work while making it easier to detect and manage deteriorating patients. While these reports are hard to correlate with our observed low usage rate, it is consistent with reports of incomplete recording of clinical parameters in obstetric EWS, especially respiratory rate (Merriel et al., 2017; Moore et al., 2019; Sheikh et al., 2017), including where EWS was implemented in well-resourced settings (Edwards et al., 2015; Lappen et al., 2010; Maguire et al., 2015). Comparatively, low usage rate of partograph has been widely reported in Nigeria (Opiah et al, 2012), and other similar settings (Mabasa S.K.M, 2018).

The success of the implementation of EWS in the intervention hospital is undoubtedly attributable to enormous support from the hospital management. Through the approval of EWS being substituted for routine vital signs charts and the dispatching of internal circulars to that effect across all obstetric wards, the major institutional barriers to implementation were broken. Additionally, the fact that the implementation was led by the local implementation team under the supervision of the local co-PI, who is a senior professor in the Obstetrics and Gynaecology department, also contributed immensely to successful implementation.

This study has some limitations. All health facilities included were tertiary hospitals that provided comprehensive emergency obstetric care services. The intervention hospital is a large university teaching hospital servicing a state, with a population of 2.37 million (National Bureau of Statistics, 2018). The feasibility and utility of implementing the EWS chart in smaller centres, including primary healthcare facilities, with weak staff strength were therefore not investigated in the present study. To improve the generalisability of findings, further multicentre studies at different levels of care including primary and secondary care hospitals are needed.

With the before-after design, it is likely that the impact of the EWS implementation on health outcomes will be stronger soon after the intervention has been implemented and that this will reduce with time. This is because there is a possibility that staff (nurses, midwives and interns) trained in each ward will not be retained within the Obstetrics department, mainly due to staff rotation. Once a critical mass of trained staff is lost from the research wards, the use of EWS will likely reduce. Although we employed surveillance and continuous training/retraining of clinical staff, low retention of trained staff remained a major limitation that significantly affected participants’ recruitment to and overall compliance with the study protocol.

## Conclusion

Findings from this research showed that the obstetric EWS can improve the quality of patient care through better monitoring frequency and medical review based on abnormally high EWS scores. The implementation was not without challenges; however, with staff education on the usefulness of EWS, provision of adequate patient monitoring equipment, coupled with continuous training and retraining of staff, EWS would potentially provide a convenient and efficient alternative patient monitoring method to cope with the unique demands faced by obstetric practice in low-resource tertiary healthcare settings

## Data Availability

Data used can be made available on reasonable request to the corresponding author

## Acknowledgement

We acknowledge the support of our colleagues, Sarah White, Hellen Allot, Alexander Manu and Hauwa Mohammed, at the Centre for Maternal and Newborn Health, Liverpool School of Tropical Medicine.

## Authors’ contributions

All authors have read and approved the manuscript. AU: conceptualisation, study design, implementation, analysis, interpretation, drafting and review. SI, IL, CC, MI: facilitated implementation, input in manuscript. MM: study design, interpretation and review, CA: study design, interpretation, drafting and review.

